# Shifting Dynamics of DENV2 in Costa Rica: Emergence of the Cosmopolitan Genotype (2024)

**DOI:** 10.1101/2025.05.28.25328475

**Authors:** Mauricio Gonzalez Elizondo, Dihala Picado Soto, Estela Cordero Laurent, Francisco Duarte Martínez, Luiz Carlos Junior Alcantara, Vagner Fonseca, Jairo Andrés Méndez Rico, Jose Lourenco, Leticia Franco, Marta Giovanetti, Claudio Soto Garita

## Abstract

Dengue remains a major public health challenge. In Costa Rica, we implemented the first nationwide genomic surveillance to track the emergence of the DENV2 Cosmopolitan genotype. Phylogenetic and eco-epidemiological analyses revealed early detection, climate-linked spread, and spatial heterogeneity. Findings underscore the need for integrated surveillance to guide adaptive responses to emerging arboviral threats.

## Text

Dengue fever, caused by the mosquito-borne dengue virus (DENV), remains a major public health threat in tropical and subtropical regions, primarily transmitted by *Aedes aegypti* (1). The rising global incidence is driven by climate change and urbanization (2). In Costa Rica, dengue transmission has become increasingly complex, with the co-circulation of all four DENV serotypes (DENV1–4), increasing the risk of co-infections and severe outcomes (3). Historically, DENV1 and DENV2 predominated, but DENV4 emerged in late 2022, followed by the reappearance of DENV3 in early 2023 after a six-year absence (2). Reported cases rose from 30,649 in 2023 (2) to 31,259 in 2024 (3), with high incidence in San José, Alajuela, and Puntarenas, prompting intensified control measures (4). Despite ongoing surveillance, significant gaps remained in understanding the genotypic shifts of dengue viruses circulating in Costa Rica. To address this, a nationwide sequencing program was launched in 2023 through Inciensa (Instituto Costarricense de Investigación y Enseñanza en Salud y Nutrición). Initial analyses identified DENV1 Genotype V, DENV2 Genotype III (Asian-American), DENV3 Genotype III, and DENV4 Genotype IIb. However, in February 2024, the DENV2 Genotype II (Cosmopolitan Genotype) was detected for the first time, marking a significant shift. By September, it had fully replaced the previously dominant Genotype III, with early cases identified in coastal districts such as Cóbano and Cahuita.

This genotype had never been reported in Costa Rica and aligns with recent detections in Peru’s Madre de Dios and Brazil’s Midwest, suggesting a broader regional pattern (5,6). Its association with more severe clinical outcomes (6) and rapid geographic spread raises concerns about increased virulence and transmissibility. Through integrated genomic, phylogenetic, and eco-epidemiological analyses, this study provides critical insights into genotype replacement dynamics, supporting the development of more adaptive and region-specific dengue control and surveillance strategies.

To further understand the ecological drivers contributing to this shift and the broader dengue transmission landscape, we analyzed recent patterns of dengue incidence in relation to climatic conditions. Over the past decade, dengue activity in Costa Rica has shown largely irregular dynamics with intermittent seasonality (2016, 2019–2020), followed by a marked increase in reported cases between 2022 and 2024 (Figure 1a). To explore the role of climate in shaping these trends, we compared monthly dengue case counts with estimated climate-driven suitability for transmission. Focusing on years with pronounced seasonal signals and high case numbers, this analysis revealed a moderate temporal correlation (r= 0.38) between monthly suitability and incidence during the 2022–2023 epidemic period. Detailed methodologies are available in the **Supplementary Appendix**.

**Figure 1.**
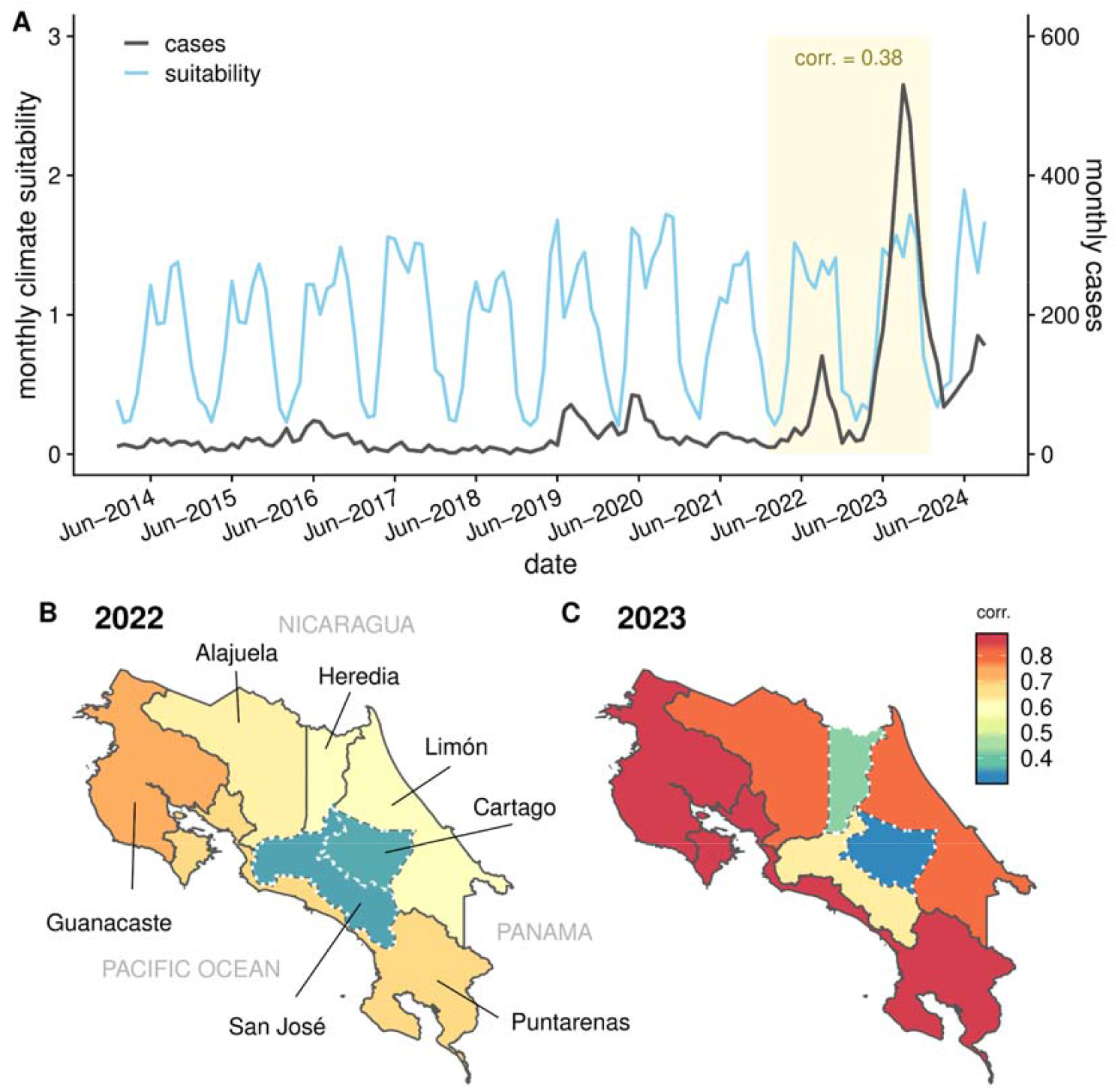
Temporal and spatial correlation between climate-driven suitability and dengue incidence in Costa Rica. a) Time series of monthly dengue cases (black line, right axis) and climate-driven suitability index for transmission (blue line, left axis) from June 2014 to November 2024. Shaded area (yellow) indicates the epidemic period during which enough cases with a clear seasonal signal are reported allowing to estimate correlation (Spearman’s test, r = 0.38, p-value<0.05) between suitability and incidence; b–c) Province-level correlation ((Spearman’s test) between monthly dengue incidence and climate suitability for the years 2022 (b) and 2023 (c). Warmer colors indicate stronger correlations. Dashed boundaries (white) mark provinces with non-significant correlation (p-value>0.05) and solid boundaries (dark grey) otherwise. In 2023, higher correlations were observed in eastern and coastal provinces where early cases of the DENV2 Cosmopolitan genotype were detected.

To further examine spatial heterogeneity, we assessed the province-level correlation between climate-driven suitability and dengue incidence across two epidemic years (2022– 2023). In 2022, correlations were intermediate-to-high and reasonably homogeneous across regions (**Figure 1b**). However, in 2023, provinces with early detection of the DENV2 Cosmopolitan genotype - such as Puntarenas and Limón - exhibited high correlations, suggesting a convergence of factors (e.g. ecological, virological, immunity) that may have facilitated localized intensification of transmission (**Figure 1c**).

Historically, DENV1 and DENV2 have been the predominant serotypes, fluctuating in their relative proportions. However, a major shift was observed in 2023–2024, characterized by the only period with co-circulation of the 4 serotypes, mirrored by a remergence of DENV3 and the emergence of DENV4 (**Figure 2a**). The remergence of DENV 3 aligns with the known ubiquitous serotype cycles observed every 7-9 years, while the emergence of DENV4 aligns with the recent expansion in South America (4). Over the years with available dengue reports, the yearly ranges in age of infection changed slightly, but no significant change could be quantified over the years (linear slope 0.17, p-value=0.048); (**Figure 2b)**. This estimate did not strongly support a significant increase in the force-of-infection over the years, as also supported by the reasonably stable estimates of climate-driven suitability (**Figure 1**), since this should be mirrored by a decreasing age of reported infections. In contrast, the age of infection increased slightly by 1.3 years for every extra circulating serotype (p-value=5.6e-06), independently of year (**Figure 2b)**, potentially reflecting that serotype mixing increases the prevalence of secondary infections, which in turn happen in already seropositive, older individuals.

**Figure 2.**
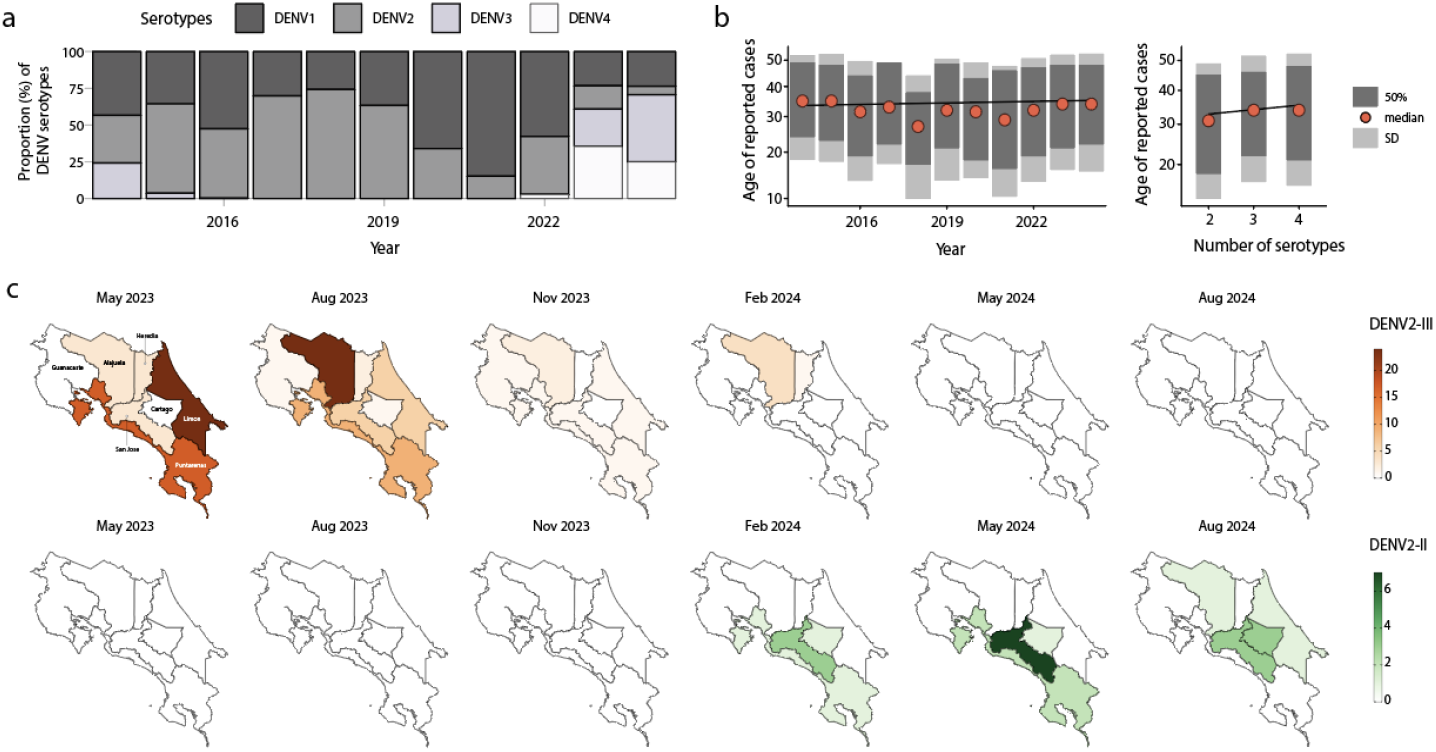
Dengue Serotype Dynamics in Costa Rica. a) Proportional distribution of DENV serotypes over time; b) Age of dengue cases over time (left) and by number of circulating serotypes (right). Median (red), interquartile range (dark gray), an standard deviation (light gray) are shown; c) Geographic distribution of DENV2 genotypes in Costa Rica from May 2023 to August 2024.

Concurrently, a marked change was observed in circulating DENV2 strains, with th previously dominant DENV2-III genotype being replaced by the DENV2-II genotype in early 2024 (**Figure 2b-c**). Between May 2023 and August 2024, the DENV2-III genotype was more prevalent, particularly in Alajuela, San José, Puntarenas, and Limón - regions historically associated with high dengue transmission. Over time, however, the DENV2-II genotype became increasingly dominant, especially in San José, Cartago, and Alajuela, while the DENV2-III genotype declined. This pattern suggests a gradual replacement, potentially driven by selective advantage, immune escape or forcing from repeated introductions from external sources.

Inciensa implemented the country’s first nationwide genomic surveillance program in 2023 as part of the epidemiological surveillance of dengue. This initiative enabled the generation of 133 whole-genome sequences (WGS) of DENV-2 from 2023/2024. Using the dynamic DENV lineage classification system (7), newly (n=110) identified DENV2 genotype III (Asian- American) genomes were classified as belonging to lineage D.1.2 and newly (n=23) identified DENV2 genotype II (Cosmopolitan) genomes were classified as belonging to lineage F.1.1.2.

DENV2 Genotype III sequences were obtained from 58 female and 52 male individuals, with a mean age of 38 years. Samples originated from Alajuela, Puntarenas, Limón, San José, Heredia, Cartago, and Guanacaste (**Table S1**). The mean genome coverage was 92.62% (range: 62.81%–97.80%) and the mean Ct value was 22 (range: 12–31). DENV2 Genotype II sequences were derived from 12 female and 11 male individuals, also with a mean age of 38 years, and were detected in Alajuela, Cartago, Limón, Puntarenas, and San José (**Table S2**). The average genome coverage was 80% (range: 77%–84%) and the mean Ct value was 24 (range: 18–28).

To gain deeper insights into the spatiotemporal dynamics of DENV2-III and the emerging dominance of DENV2-II, we performed phylodynamic analyses (**Figure 3**), which revealed a well-supported monophyletic clade of Costa Rican DENV2-III, suggesting that local persistence has been sustained after introduction events from Central America over the last decade (**Figure 3a**).

**Figure 3.**
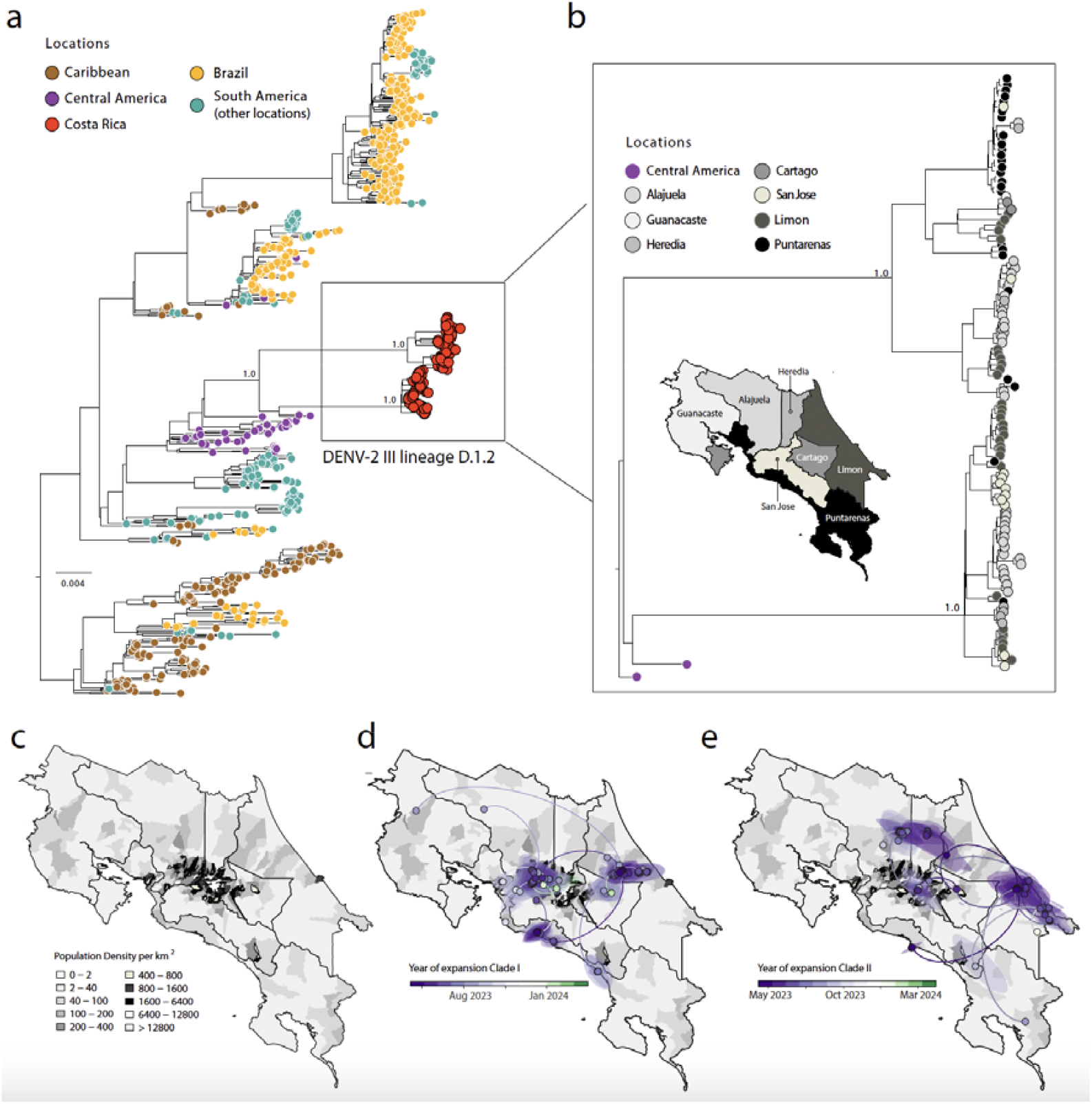
Evolutionary and spatiotemporal expansion of DENV2-III in Costa Rica. a) Maximum likelihood (ML) phylogeny of DENV2-III, showing Costa Rican sequences (red) within the broader Central and South American clade. Phylogenetic support is indicated at key nodes; b) ML reconstruction of DENV2-III lineage D.1.2 dispersal within Costa Rica; c) Population density map of Costa Rica, highlighting areas with elevated transmission potential; d, e) Spatiotemporal expansion of DENV-2 III lineage D.1.2, illustrating the progressive spread of two distinct clades—Clade I and II—from urban centers to coastal regions between May 2023 and March 2024.

Maximum likelihood phylogenetic reconstruction revealed the co-circulation of two distinct clades within the DENV-2 III lineage D.1.2 i referred to here as clade I and II (**Figure 3b**). Although these clades are phylogenetically distinct, both belong to the same lineage. Phylogeographic analysis further elucidated patterns of viral spread across regions. Initial circulation was concentrated in Alajuela, Cartago, and San José, followed by expansion toward Puntarenas and Limón coastal regions. Clade I (**Figure 3d**) was estimated to have emerged in May 2023, with a 95% highest posterior density (HPD) interval ranging from April to late May 2023. Its inferred transmission pathways extended from San José and Cartago toward Puntarenas and Limón by early 2024. Clade II (**Figure 3e**) was detected as early as June 2023, with a 95% HPD covering a similar timeframe, and exhibited a broader dispersal pattern encompassing the densely populated areas of Alajuela and San José. The spatial overlap of these sublineages with regions of high population density (**Figure 3c**) underscores the role of urban centers as key transmission hubs facilitating the spread of DENV-2.

Further phylogenetic resolution of DENV2-II sequences (**Figure 4**) revealed a distinct evolutionary trajectory compared to DENV2-III, supporting the hypothesis of a recent introduction followed by rapid establishment in Costa Rica. The time stamped phylogenetic tree indicated that at least two independent introduction of the DENV2-II F.1.1.2 lineage likely occurred, potentially mediated by regional viral flow from Latin American countries, including Bolivia and Brazil and resulted in the establishment of a well-supported monophyletic clade. Bayesian time-scaled phylogenetic analysis of this clade suggests an emergence around October 2023, with a 95% HPD interval spanning from September to late November 2023. Early circulation was primarily concentrated in Puntarenas, Limón, and Cartago, with subsequent dissemination into San José, Alajuela, and Heredia (**Figure 4b**).

**Figure 4.**
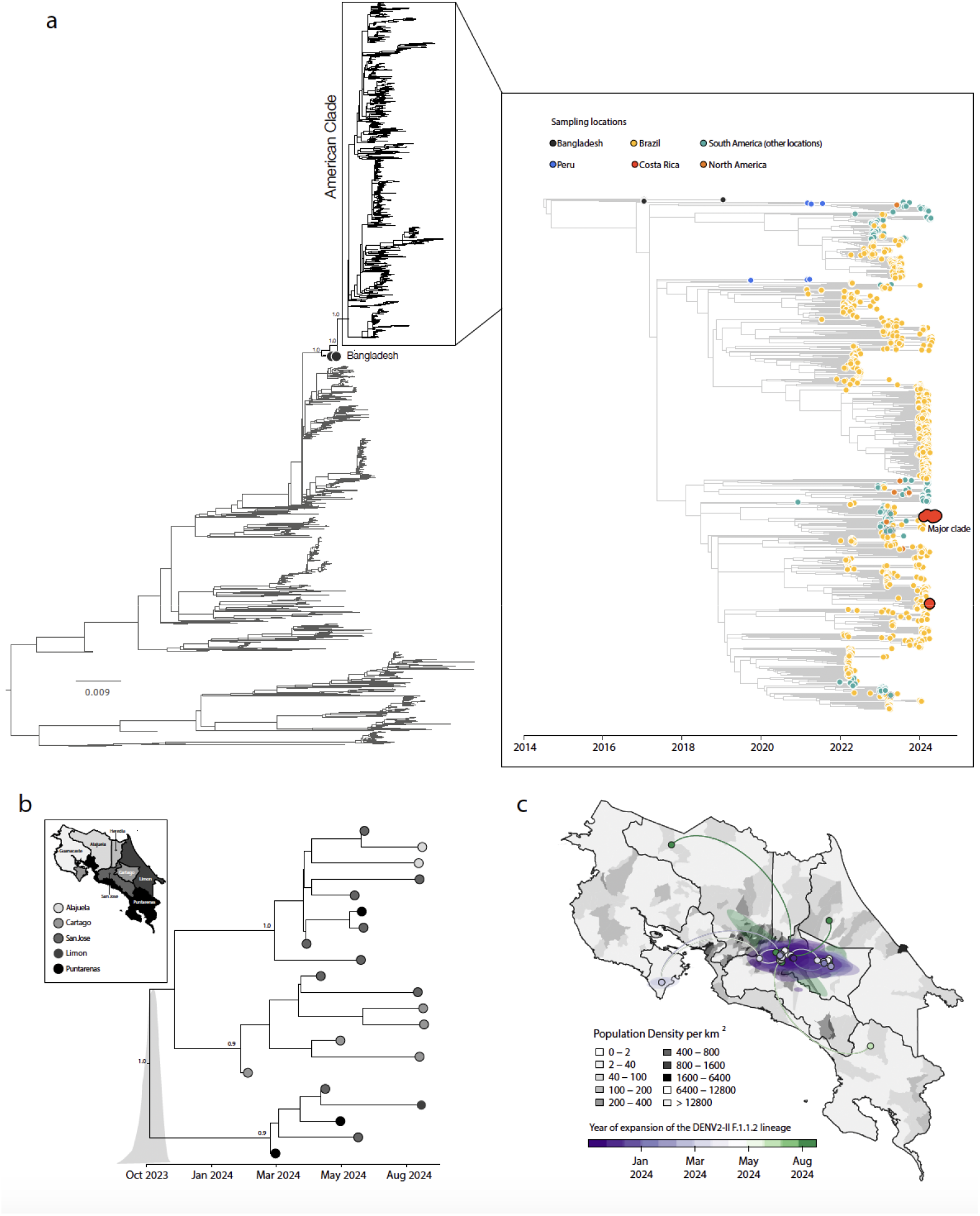
Emergence and spread of DENV2-II in Costa Rica. a) Maximum likelihood (ML) phylogeny of DENV2-II, highlighting the American clade. The inset on the right displays the time-stamped phylogenetic tree, showing two independent introductions of this genotype into Costa Rica, including one major transmission cluster. Tips are colored according to samplin location; b) Time-scaled phylogenetic reconstruction of the major DENV2-II clade, illustrating its expansion from Puntarenas, Limón, and Cartago in early 2024; c) Spatiotemporal dynamics of the major DENV2-II clade in Costa Rica.

Reconstruction of viral dispersal for this major clade (**Figure 4c)** further highlighted its rapid establishment across densely populated areas. Initially detected in coastal and central provinces, the virus quickly spread into high-transmission hubs, particularly those characterized by high population density.

## Conclusion

This study offers the first nationwide genomic and eco-epidemiological assessment of dengue in Costa Rica, highlighting the recent replacement of DENV2-III (Asian-American) by the emerging DENV2-II (Cosmopolitan) genotype. Using whole-genome sequencing, phylodynamics, and climate-based modeling, we demonstrate how viral evolution and ecological factors shape transmission. Similar genotype replacements have occurred elsewhere, often linked to immunity shifts or viral fitness advantages (8–10). However, we found no evidence of increased transmission due to climate or age-related shifts in infection. DENV2-II, first detected in early 2024, rapidly replaced DENV2-III despite a decline in overall DENV2 circulation (15.8% in 2023 to 9.3% in 2024). Importantly, this shift was not associated with increased case severity, and no deaths in 2024 were attributed to this genotype. At least two independent introduction events led to widespread dissemination across provinces, consistent with patterns observed in Brazil and Southeast Asia (11, 12). The Cosmopolitan genotype, now dominant worldwide, was previously reported in Peru (2019), Brazil (2022), Colombia (2023), and Costa Rica (2024) (5,6,13). A moderate correlation (r = 0.38) between climate suitability and dengue incidence, especially in early DENV2-II hotspots, supports the role of environmental conditions in amplifying transmission (2,14). Its spread from coastal regions to urban centers such as San José and Alajuela reflects the influence of urbanization and mobility over ecological barriers

(15–17). While our genomic data are substantial, uneven geographic sampling and reliance on passive surveillance limited detection in some areas. Overall, this study highlights how genotype introductions, ecological conditions, and human mobility interact to shape dengue transmission dynamics in Costa Rica. Strengthening real-time genomic surveillance systems—integrated with environmental and mobility data—is essential for early detection of emerging threats and for guiding timely, targeted public health interventions.

## Data Availability

All generated data have been deposited in GenBank and are available under accession numbers PV628991 to PV629123.

## Acknowledgement

This study was supported by PAHO, USAID, and in part by the National Institutes of Health (NIH) USA grant U01 AI151698 for the United World Arbovirus Research Network (UWARN) and the and the Novo Nordisk Foundation (NNF24OC0094346). The authors would also like to acknowledge the Global Consortium to Identify and Control Epidemics – CLIMADE (T.O., L.C.J.A., E.C.H., J.L., M.G.) (https://climade.health/).

## Author contributions

Conception and design: MGE, LF, and MG; Investigations: MGE, DPS, ECL, FDM, LCJA, VF, JMR, JL, LF, MR and MSG; Data Analysis: MGE, DPS, ECL, FDM, LCJA, VF, JMR, JL, LF, MR and MSG; Visualization: VF, JL and MG; Writing – Original: MGE, JL and MG; Revision: MGE, DPS, ECL, FDM, LCJA, VF, JMR, JL, LF, MR and MSG; Data Analysis: MGE, DPS, ECL, FDM, LCJA, VF, JMR, JL, LF, MR and MSG.

## References

1. Pan-American Health Organization (PAHO), 2025. https://www.paho.org/en/documents/dengue-epidemiological-situation-region-americas-epidemiological-week-08-2025

2. Nakase, T., Giovanetti, M., Obolski, U. et al. Population at risk of dengue virus transmission has increased due to coupled climate factors and population growth. Commun Earth Environ 5, 475 (2024). 10.1038/s43247-024-01639-6

3. Ministerio de Salud Costa Rica. https://www.ministeriodesalud.go.cr/

4. Pan-American Health Organization (PAHO), Dengue cases. https://www3.paho.org/data/index.php/en/mnu-topics/indicadores-dengue-en/dengue-regional-en/315-reg-dengue-incidence-en.html

5. García MP, Padilla C, Figueroa D, et al. Emergence of the Cosmopolitan genotype of dengue virus serotype 2 (DENV2) in Madre de Dios, Peru, 2019. Rev Peru Med Exp Salud Publica. 2022 Jan-Mar;39(1):126–128. doi: 10.17843/rpmesp.2022.391.10861.

6. Giovanetti M, Pereira LA, Santiago GA, et al. Emergence of Dengue Virus Serotype 2 Cosmopolitan Genotype, Brazil. Emerg Infect Dis. 2022 Aug;28(8):1725–1727. doi: 10.3201/eid2808.220550.

7. Hill V, Cleemput S, Pereira JS, et al. A new lineage nomenclature to aid genomic surveillance of dengue virus. PLoS Biol. 2024 Sep 16;22(9):e3002834. doi: 10.1371/journal.pbio.3002834.

8. Holmes EC, Twiddy SS. The origin, emergence and evolutionary genetics of dengue virus. Infect Genet Evol. 2003 May;3(1):19–28. doi: 10.1016/s1567-1348(03)00004-2.

9. Katzelnick LC, Gresh L, Halloran ME, et al. Antibody-dependent enhancement of severe dengue disease in humans. Science. 2017 Nov 17;358(6365):929–932. doi: 10.1126/science.aan6836.

10. Messina JP, Brady OJ, Golding N, et al. The current and future global distribution and population at risk of dengue. Nat Microbiol. 2019 Sep;4(9):1508–1515. doi: 10.1038/s41564-019-0476-8.

11. Yu, H., Kong, Q., Wang, J. et al. Multiple Lineages of Dengue Virus Serotype 2 Cosmopolitan Genotype Caused a Local Dengue Outbreak in Hangzhou, Zhejiang Province, China, in 2017. Sci Rep 9, 7345 (2019). 10.1038/s41598-019-43560-5

12. Colón-González, F.J., Gibb, R., Khan, K. et al. Projecting the future incidence and burden of dengue in Southeast Asia. Nat Commun 14, 5439 (2023). 10.1038/s41467-023-41017-y

13. INS, Colombia, 2023. https://www.ins.gov.co/BibliotecaDigital/comunicado-tecnico-identificacion-de-la-circulacion-genotipo-cosmopolitan-del-virus-del-dengue-2-en-colombia.pdf

14. Salje H, Lessler J, Maljkovic Berry I, et al. Dengue diversity across spatial and temporal scales: Local structure and the effect of host population size. Science. 2017 Mar 24;355(6331):1302–1306. doi: 10.1126/science.aaj9384.

15. Kiang MV, Santillana M, Chen JT, et al. Incorporating human mobility data improves forecasts of Dengue fever in Thailand. Sci Rep 11, 923 (2021). 10.1038/s41598-020-794380

16. Gubler DJ. Dengue, Urbanization and Globalization: The Unholy Trinity of the 21(st) Century. Trop Med Health. 2011 Dec;39(4 Suppl):3–11. doi: 10.2149/tmh.2011-S05.

17. Grubaugh, N.D., Ladner, J.T., Lemey, P. et al. Tracking virus outbreaks in the twenty-first century. Nat Microbiol 4, 10–19 (2019). 10.1038/s41564-018-0296-2.

