## Appendix for "Shifting Dynamics of DENV2 in Costa Rica: Emergence of the Cosmopolitan Genotype (2024)"

**Supplementary Material**

**Material and Methods**

***Ethics statement***

The Pan American Health Organization Ethics Review Committee (PAHOERC) reviewed and approved this project (Ref. No. PAHO-2024-08-0029). The samples used in this study were analyzed as part of the routine epidemiological surveillance of arboviruses at the Virology National Reference Center (CNRV, in Spanish) at Inciensa, which is the official laboratory of the Costa Rican Ministry of Health.

***Sample collection and whole genome sequencing***

Samples were collected from patients exhibiting clinical symptoms consistent with dengue virus infection. Nucleic acid extraction was performed using the QIAamp Viral RNA Mini Kit (Qiagen). Subsequently, the extracted RNA was subjected to real-time reverse transcription PCR (RT-qPCR) targeting DENV serotypes 1–4, as previously described (1). Samples that tested positive (n = 133) and presented a cycle threshold (Ct) value of ≤30 were selected for whole-genome amplification using the CDC Next Generation Sequencing Protocol for DENV 1–4 with the Illumina MiSeq platform (2). This protocol, developed by the CDC, was transferred to the Arbovirus Diagnosis Laboratory Network of the Americas (RELDA) through the VIGENDA program coordinated by the Pan-American Health Organization (PAHO). Library preparation was carried out using the COVIDseq Kit (Illumina, San Diego, USA), originally developed for SARS-CoV-2 genomic studies but subsequently adapted for other viral targets (3, 4). Sequencing was then conducted on the Illumina MiSeq platform (Illumina, San Diego, USA), following the manufacturer’s recommendations, utilizing V2 cartridges with a 2 × 150 cycles paired-end run. Adapter trimming and quality filtering of the raw reads were performed using Trim-Galore v0.6.5 (5) with default parameters for Illumina paired-end reads. Processed reads were mapped to the reference genome using BWA-MEM v0.7.17-4 (6) with default settings. The reference genome used for alignment was the serotype-specific DENV2_Reference_Genome_PR1994, provided by the CDC’s Dengue Branch in Puerto Rico. Primer sequences were removed using iVar v1.3.1 (7). Consensus sequences were then generated from the aligned reads using Samtools v1.10-13 (8) and iVar consensus v1.3.1, applying a minimum coverage depth threshold of 10 reads and the default frequency threshold for consensus base calling. Initial genotyping was performed using the Flavivirus Genotyping Tool (Version 0.1; https://www.rivm.nl/mpf/typingtool/flavivirus/). Subsequently, lineage classification was conducted using either the Genome Detective Dengue Typing Tool or Nextclade (9, 10), employing the DENV-2 reference dataset for lineage determination.

***Phylogenetic and phylodynamic inferences***

We constructed phylogenetic trees to investigate the evolutionary relationship of sequenced DENV2 genomes from Costa Rica (DENV2-III n=110 and DENV2-II n=23) in comparison to globally sampled sequences (n=654 and n=2213 respectively). Sequence alignment was performed using MAFFT (11) and manually curated in AliView (12). Preliminary maximum likelihood phylogenies were reconstructed using IQ-TREE 2 under the HKY+G4 substitution model (13). Time-scaled phylogenies were inferred with TreeTime (14), while Bayesian phylogenetic analyses were conducted using BEAST (15). To ensure the robustness of the temporal framework, TempEst (16) was employed to assess the presence of a temporal signal. For Bayesian inference, we implemented a rigorous model selection approach, utilizing both path-sampling (PS) and stepping-stone (SS) methods to identify the most appropriate molecular clock model (17). The uncorrelated relaxed molecular clock was selected for all datasets based on marginal likelihood estimation, in combination with the codon-based SRD06 nucleotide substitution model and the Bayesian Skyline coalescent model. To reconstruct the geographic dissemination of the identified 2022-2023 transmission clade, we employed a relaxed random walk diffusion model (18, 19), which accommodates heterogeneity in dispersal rates among branches, incorporating a Cauchy distribution and a jitter window site of 0.01 (20). Each sequence was georeferenced with latitude and longitude coordinates to enable spatiotemporal analyses. Bayesian phylogenetic inference was performed using BEAST v1.10.4, with two independent Markov Chain Monte Carlo chains (MCMC) runs of 50 million interactions, sampling every 10,000 steps. Convergence was assessed in Tracer, ensuring effective sample size (ESS) >200 for all key parameters. Maximum clade credibility (MCC) trees were summarized using TreeAnnotator after discarding the initial 10% os samples as burn-in. Finally, we employed the R package 'seraphim' version 1.0 (21) to extract and visualize spatiotemporal patterns embedded within the posterior tree distribution.

***Eco-epidemiological modelling***

The epidemiological data on weekly confirmed cases of DENV in Costa Rica, from 2013 to 2024, were obtained from the CNRV and aggregated at a monthly level. We calculated the theoretical climate-based transmission potential of the dengue virus using the following mathematical expression (index P), where u stands for humidity and t for temperature:

$P_{(u,t)}=\frac{{a_{(u)}^{v}}^{2}\phi_{(t)}^{v\to h}\phi_{(t)}^{h\to v}\gamma_{\left( t \right)}^{v}\gamma^{h}}{\mu_{\left( u,t \right)}^{v}(\sigma^{h}+\mu^{h})(\gamma^{h}+\mu_{\left( u,t \right)}^{v})}$

(22)

Briefly, the index uses mathematical expressions of empirically demonstrated relationships between DENV and *Ae. aegypti* traits and meteorological variables. Climate-dependent traits include the extrinsic incubation period ($\gamma_{\left( t \right)}^{v}$), adult mosquito lifespan ($\mu_{\left( u,t \right)}^{v}$), adult mosquito biting rate ($a_{(u)}^{v}$transmission probability per mosquito bite from infected human to susceptible mosquito ($\phi_{(t)}^{h\to v}$) and from infected mosquito to susceptible human ($\phi_{(t)}^{v\to h}$). Traits that are climate-independent include intrinsic incubation period ($\gamma^{h}$), human lifespan ($\mu^{h}$) and human infectious period ($\sigma^{h}$), which are calibrated from published reports. Full methodological details, technical validation and parameterization of Index P can be found in Nakase et al (22). Monthly climate data for Costa Rica was obtained from Copernicus.eu satellite climate data (23).

**References**

1. Santiago GA, Vergne E, Quiles Y, et al. Analytical and clinical performance of the CDC real time RT-PCR as-say for detection and typing of dengue virus. PLoS Negl Trop Dis. 2013; 11;7(7):e2311.

2. Vogels CBF, Hill V, Breban MI, Chaguza C, Paul LM, Sodeinde A, Taylor-Salmon E, Ott IM, Petrone ME, Dijk D, Jonges M, Welkers MRA, Locksmith T, Dong Y, Tarigopula N, Tekin O, Schmedes S, Bunch S, Cano N, Jaber R, Panzera C, Stryker I, Vergara J, Zimler R, Kopp E, Heberlein L, Herzog KS, Fauver JR, Morrison AM, Michael SF, Grubaugh ND. DengueSeq: a pan-serotype whole genome amplicon sequencing protocol for dengue virus. BMC Genomics. 2024 May 1;25(1):433. doi: 10.1186/s12864-024-10350-x. PMID: 38693476; PMCID: PMC11062901.

3. Guimarães NR, Tomé LR, Lamounier LO, et al. Genomic Surveillance of Monkeypox Virus, Minas Gerais, Brazil, 2022. Emerg Infect Dis. 2023;29(6):1270-1273. https://doi.org/10.3201/eid2906.230113

4. Chen NFG, Chaguza C, Gagne L, et al. Development of an amplicon-based sequencing approach in response to the global emergence of mpox. PLoS Biol. 2023 Jun 13;21(6):e3002151. doi: 10.1371/journal.pbio.3002151.

5.Krueger, F. (2012). Trim Galore. https://www.bioinformatics.babraham.ac.uk/projects/trim_galore/

6. Li H, Durbin R. Fast and accurate short read alignment with Burrows-Wheeler transform. Bioinformatics. 2009 Jul 15;25(14):1754-60. doi: 10.1093/bioinformatics/btp324. Epub 2009 May 18. PMID: 19451168; PMCID: PMC2705234.

7. Grubaugh, N. D., Gangavarapu, K., Quick, J., Matteson, N. L., De Jesus, J. G., Main, B. J., Tan, A. L., Paul, L. M., Brackney, D. E., Grewal, S., Gurfield, N., Van Rompay, K. K. A., Isern, S., Michael, S. F., Coffey, L. L., Loman, N. J., & Andersen, K. G. (2019). An amplicon-based sequencing framework for accurately measuring intrahost virus diversity using PrimalSeq and iVar. Genome Biology, 20(1), 8. https://doi.org/10.1186/s13059-018-1618-7

8. Danecek, P., Bonfield, J. K., Liddle, J., Marshall, J., Ohan, V., Pollard, M. O., Whitwham, A., Keane, T., McCarthy, S. A., Davies, R. M., & Li, H. (2021). Twelve years of SAMtools and BCFtools. GigaScience, 10(2), giab008. https://doi.org/10.1093/gigascience/giab008

9. Vilsker M, Moosa Y, Nooij S, Fonseca V, Ghysens Y, Dumon K, Pauwels R, Alcantara LC, Vanden Eynden E, Vandamme AM, Deforche K, de Oliveira T. Genome Detective: an automated system for virus identification from high-throughput sequencing data. Bioinformatics. 2019 Mar 1;35(5):871-873. doi: 10.1093/bioinformatics/bty695. PMID: 30124794; PMCID: PMC6524403.

10. Aksamentov, I., Roemer, C., Hodcroft, E. B., & Neher, R. A., (2021). Nextclade: clade assignment, mutation calling and quality control for viral genomes. Journal of Open Source Software, 6(67), 3773, https://doi.org/10.21105/joss.03773

11. Katoh K, Rozewicki J, Yamada KD. MAFFT online service: multiple sequence alignment, interac-tive sequence choice and visualization. Brief Bioinform. 2019; 19;20(4):1160-1166.

12. Larsson A. AliView: a fast and lightweight alignment viewer and editor for large datasets. Bio-informatics. 2014; 30:3276–8.

13. Nguyen L-T, Schmidt HA, von Haeseler A, Minh BQ. IQ-TREE: a fast and effective stochastic al-gorithm for estimating maximum-likelihood phylogenies. Mol Biol Evol. 2015;32(1):268–74.

14. Sagulenko P, Puller V, Neher RA. TreeTime: Maximum-likelihood phylodynamic analysis. Virus Evol. 2018 Jan 8;4(1):vex042. doi: 10.1093/ve/vex042.

15. Suchard MA, Lemey P, Baele G, Ayres DL, Drummond AJ, Rambaut A. Bayesian phylogenetic and phylodynamic data integration using BEAST 1.10. Virus Evol. 2018 Jun 8;4(1):vey016. doi: 10.1093/ve/vey016.

16. Rambaut A, Lam TT, Max Carvalho L, Pybus OG. Exploring the temporal structure of heterochronous sequences using TempEst (formerly Path-O-Gen). Virus Evol. 2016 Apr 9;2(1):vew007. doi: 10.1093/ve/vew007.

17. Baele G, Li WL, Drummond AJ, Suchard MA, Lemey P. Accurate model selection of relaxed molecular clocks in bayesian phylogenetics. Mol Biol Evol. 2013 Feb;30(2):239-43. doi: 10.1093/molbev/mss243. Epub 2012 Oct 22.

18. Lemey P, Rambaut A, Welch JJ, Suchard MA. Phylogeography takes a relaxed random walk in continuous space and time. Mol Biol Evol. 2010 Aug;27(8):1877-85. doi: 10.1093/molbev/msq067. Epub 2010 Mar 4.

19. Pybus OG, Suchard MA, Lemey P, et al. Unifying the spatial epidemiology and molecular evolution of emerging epidemics. Proc Natl Acad Sci U S A. 2012 Sep 11;109(37):15066-71. doi: 10.1073/pnas.1206598109.

20. Dellicour S, Gill MS, Faria NR, et al. Relax, Keep Walking - A Practical Guide to Continuous Phylogeographic Inference with BEAST. Mol Biol Evol. 2021 Jul 29;38(8):3486-3493. doi: 10.1093/molbev/msab031.

21. Dellicour S, Rose R, Faria NR, et al. SERAPHIM: studying environmental rasters and phylogenetically informed movements. Bioinformatics. 2016 Oct 15;32(20):3204-3206. doi: 10.1093/bioinformatics/btw384.

22. Nakase, T., Giovanetti, M., Obolski, U. et al. Population at risk of dengue virus transmission has increased due to coupled climate factors and population growth. Commun Earth Environ 5, 475 (2024). https://doi.org/10.1038/s43247-024-01639-6

23. Copernicus Climate Data Store. Essential climate variables for assessment of climate variability from 1979 to present, <https://cds.climate.copernicus.eu/cdsapp#!/dataset/ecv-for-climate-change>.
